# COVID-19: age, Interleukin-6, C-Reactive Protein and lymphocytes as key clues from a multicentre retrospective study in Spain

**DOI:** 10.1101/2020.05.13.20101345

**Authors:** Aurora Jurado, María C. Martín, Cristina Abad-Molina, Antonio Orduña, Alba Martínez, Esther Ocaña, Oscar Yarce, Ana M. Navas, Antonio Trujillo, Luis Fernández, Esther Vergara, Beatriz Rodríguez, Bibiana Quirant, Eva Martínez-Cáceres, Manuel Hernández, Janire Perurena-Prieto, Juana Gil, Sergi Cantenys, Gema González-Martínez, María T. Martínez-Saavedra, Ricardo Rojo, Francisco M. Marco, Sergio Mora, Jesús Ontañón, Marcos López-Hoyos, Gonzalo Ocejo-Vinyals, Josefa Melero, Marta Aguilar, Delia Almeida, Silvia Medina, María C. Vegas, Yesenia Jiménez, Álvaro Prada, David Monzón, Francisco Boix, Vanesa Cunill, Juan Molina

## Abstract

**Background:** SARS-CoV-2 infection has widely spread to the hugest public health challenge to date, COVID-19 pandemic. Different fatality rates among countries are probably due to non-standardized records being carried out by local health authorities. Spanish case-fatality rate is 11.94%, far higher to those reported in Asia or by other European countries. A multicenter retrospective study was performed of demographic, clinical, laboratory and immunological features of 574 Spanish COVID-19 hospitalized patients and their outcomes. The use of use of renin-angiotensin system blockers was also analyzed as a risk factor.

**Results:** In this study, 27.7% of cases presented a mild curse, 42% a moderate one and for 30.3% of cases, the course was severe. Ages ranged from 18 to 98 (average 63.2). Fifty eight percent (58.9%) of patients were male. Interleukin 6 was higher as severity increased. On the other hand, CD8 lymphocyte count was significantly lower as severity grew and subpopulations CD4, CD8, CD19 and NK showed concordant lowering trends. Severity-related natural killer percent descents were evidenced just within aged cases. A significant severity-related decrease of CD4 lymphocytes was found in males. The use of renin-angiotensin system blockers was associated with moderate or mild disease courses.

**Conclusions:** Age and age-related comorbidities, such as dyslipidaemia, hypertension or diabetes, determined more frequent severe forms of the disease in this study than in previous literature cohorts. Our cases are older than those so far reported and clinical course of the disease is found to be impaired by age. Immunosenescence might be therefore a suitable explanation for immune system effectors severity-related hampering. Adaptive immunity would go exhausted and a huge ineffective and almost deleterious innate response would account for COVID-19 severity. Renin-angiotensin system blockers treatment in hypertensive patients has a protective effect as regarding COVID-19 severity.

## BACKGROUND

SARS-CoV-2 infection has become worldwide widespread. Never before did we experience a health emergency like this. At the time of writing, four months after the first diagnosed case [1] the virus has infected 4 690 287 people, with an overall case-fatality rate of 6.69% [2] far exceeding the 1% reported outside the epicentre by early studies [3]. It can be traced back to the end of February, when the pandemic started to rapidly expand, hitting the hardest some European countries, such as Spain, with case-fatality rates around 11.94%. So far, we lack an explanation to this particular behaviour, although it could be due either to different local approaches for records and statistics of infected patients within countries or to demographical features. Absolute mortality rates are far higher in Spain than those reported in Asia or other European countries [4].

It was by 2002, in the SARS-CoV epidemic, when a coronavirus was for the first time revealed to be highly pathogenic. Coronaviruses were until then considered to cause only mild infections, mainly in immunocompromised people [5]. Compared with SARS-CoV, SARS-CoV-2 has shown a much higher infectivity with a doubling time of 2.3-3.3 days, and a basic reproductive number [R_0_] of 5.7 [6]. SARS-CoV-2 can be considered specially challenging due to several intrinsic and extrinsic characteristics. It has a highly variable prevalence and outcomes within countries depending on age, weather and social habits.

Angiotensin-converting enzyme 2 (ACE2) is the receptor for SARS-CoV-2 and plays a key role in human infection [7], ACE2 has two isoforms; a large one anchored to the cell membrane [8] and a small soluble isoform lacking anchorage to the membrane and circulating in low amounts in peripheral blood [9]. Hence, it has been suggested that the use of drugs that increase the expression of ACE2, such as angiotensin converting enzyme inhibitors and angiotensin II receptor blockers, could enhance infection [10]. On the other hand, increasing soluble ACE2 may be a therapeutic tool to block the virus [11]. The habit of smoking can as well rise ACE2 expression and might be therefore a risk factor for SARS CoV2 infection [12].

Both innate and adaptive responses are involved in SARS-CoV-2 fighting [13]. An accurate immune response is essential for infection resolution. In SARS-CoV-2 infection, an aberrant immune response itself might be the key to understand the immunopathogenesis. It looks as if the progression to severe COVID-19 were associated with a poor adaptive immune response [14] and with an exacerbation of the innate immune response, with an increase in plasma levels of cytokines and pro-inflammatory chemokines [15].

Understanding the pathogenesis of the virus as well as identifying risk or severity factors for COVID-19, are key points for identifying disease evolution biomarkers and taking immediate preventive actions.

The aim of this study was therefore to obtain, within the shortest possible time, a reliable snapshot of the demographic and clinical characteristics of COVID-19 patients admitted to Spanish hospitals along the first month of the pandemic and to reveal risk factors regarding outcome severity. This knowledge would help manage both clinical and public health decisions.

## RESULTS

A total of 574 SARS-CoV-2 infected inpatients from 19 Spanish Hospitals were included. Twenty-seven percent (27.7%) of cases presented a mild curse, 42.0% a moderate one and 30.3% a severe one. By data collection deadline, 266 patients had been discharged and 84 had died. Descriptive baseline characteristics of population (valid n, frequencies, percentages, mean, median, standard deviation and interquartile range) are shown in Table 1. Categorical variables stratified by severity are shown in Table 2.

**Table 1.**
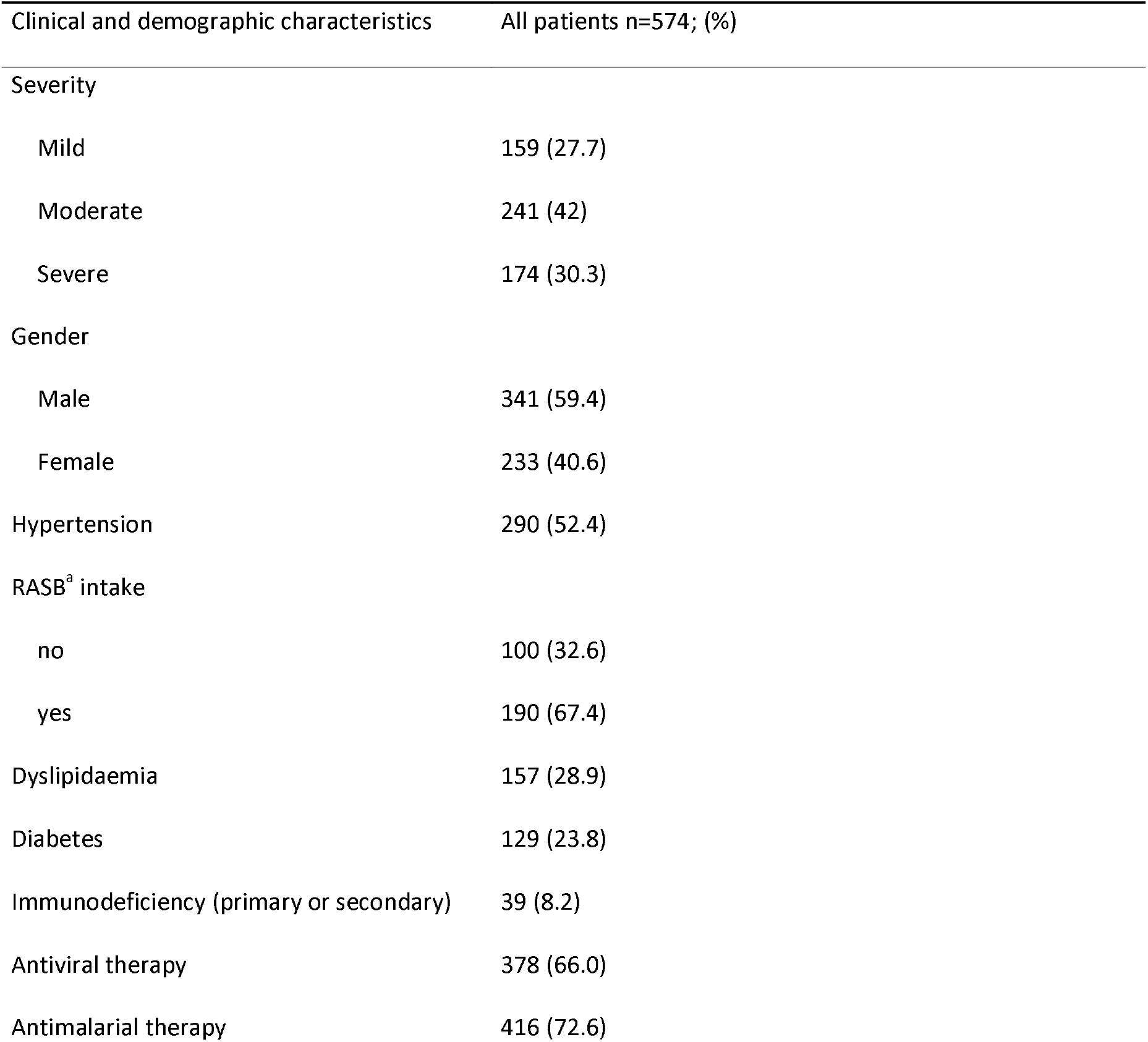
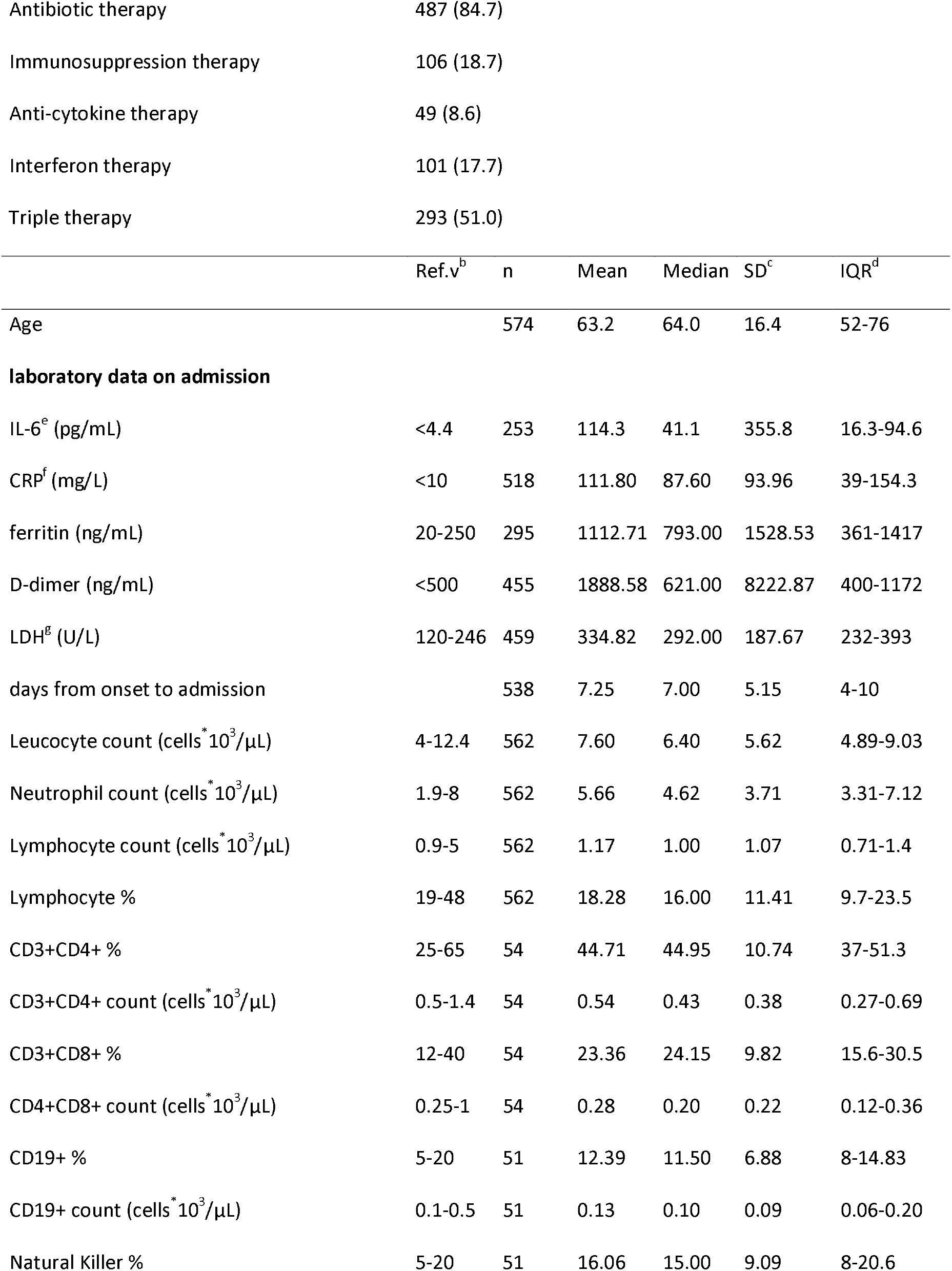
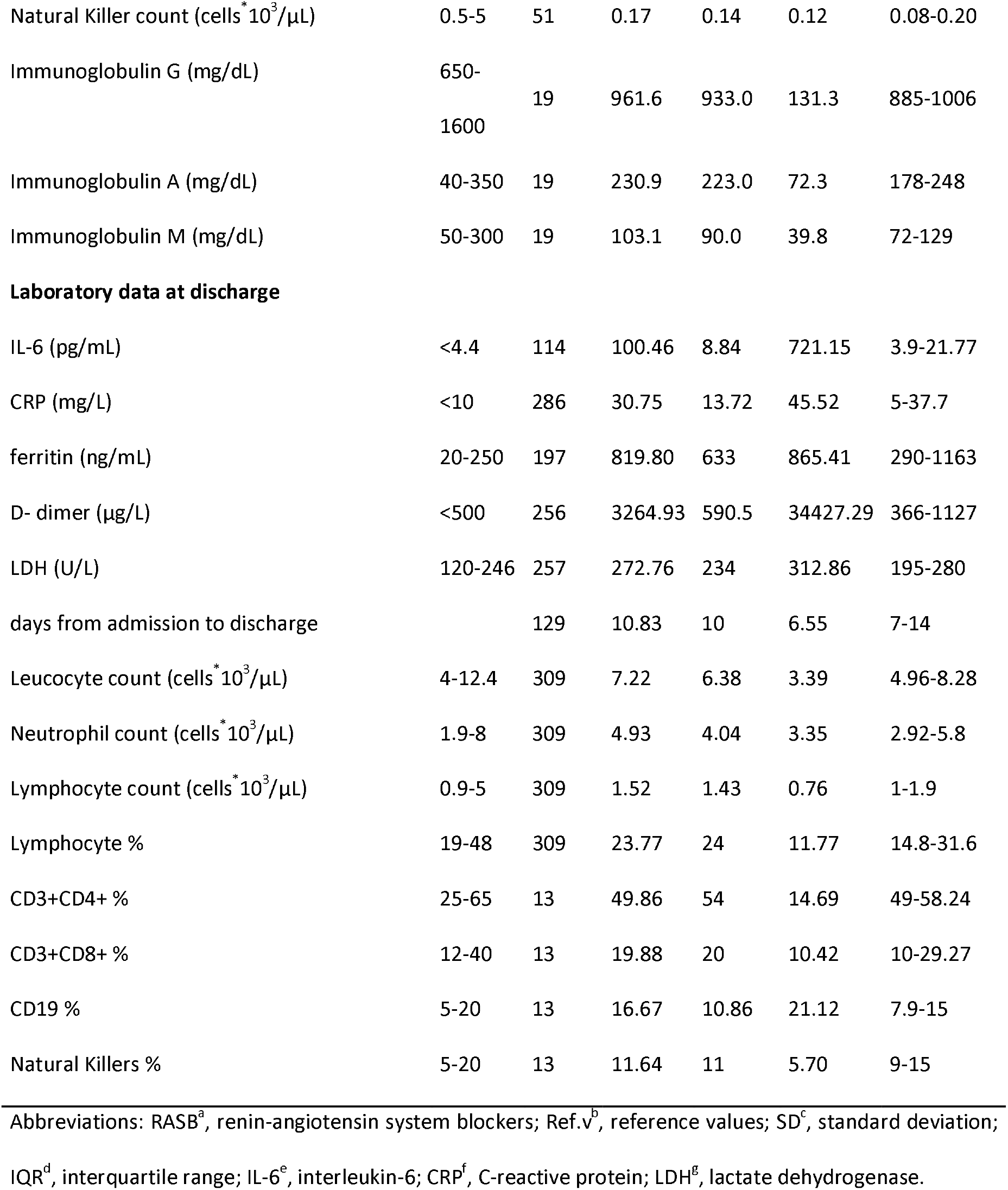
Baseline characteristics of the study population.

| Clinical and demographic characteristics | All patients n=574; (%) |
| --- | --- |
| Severity |  |
| Mild | 159 (27.7) |
| Moderate | 241 (42) |
| Severe | 174 (30.3) |
| Gender |  |
| Male | 341 (59.4) |
| Female | 233 (40.6) |
| Hypertension | 290 (52.4) |
| RASB<sup>a</sup> intake |  |
| no | 100 (32.6) |
| yes | 190 (67.4) |
| Dyslipidaemia | 157 (28.9) |
| Diabetes | 129 (23.8) |
| Immunodeficiency (primary or secondary) | 39 (8.2) |
| Antiviral therapy | 378 (66.0) |
| Antimalarial therapy | 416 (72.6) |

| Antibiotic therapy | 487 (84.7) |  |  |  |  |  |
| --- | --- | --- | --- | --- | --- | --- |
| Immunosuppression therapy | 106 (18.7) |  |  |  |  |  |
| Anti-cytokine therapy | 49 (8.6) |  |  |  |  |  |
| Interferon therapy | 101 (17.7) |  |  |  |  |  |
| Triple therapy | 293 (51.0) |  |  |  |  |  |
|  | Ref.v<sup>b</sup> | n | Mean | Median | SD<sup>c</sup> | IQR<sup>d</sup> |
| Age |  | 574 | 63.2 | 64.0 | 16.4 | 52-76 |
| **laboratory data on admission** |  |  |  |  |  |  |
| IL-6<sup>e</sup> (pg/mL) | <4.4 | 253 | 114.3 | 41.1 | 355.8 | 16.3-94.6 |
| CRP<sup>f</sup> (mg/L) | <10 | 518 | 111.80 | 87.60 | 93.96 | 39-154.3 |
| ferritin (ng/mL) | 20-250 | 295 | 1112.71 | 793.00 | 1528.53 | 361-1417 |
| D-dimer (ng/mL) | <500 | 455 | 1888.58 | 621.00 | 8222.87 | 400-1172 |
| LDH<sup>g</sup> (U/L) | 120-246 | 459 | 334.82 | 292.00 | 187.67 | 232-393 |
| days from onset to admission |  | 538 | 7.25 | 7.00 | 5.15 | 4-10 |
| Leucocyte count (cells*10<sup>3</sup>/μL) | 4-12.4 | 562 | 7.60 | 6.40 | 5.62 | 4.89-9.03 |
| Neutrophil count (cells*10<sup>3</sup>/μL) | 1.9-8 | 562 | 5.66 | 4.62 | 3.71 | 3.31-7.12 |
| Lymphocyte count (cells*10<sup>3</sup>/μL) | 0.9-5 | 562 | 1.17 | 1.00 | 1.07 | 0.71-1.4 |
| Lymphocyte % | 19-48 | 562 | 18.28 | 16.00 | 11.41 | 9.7-23.5 |
| CD3+CD4+ % | 25-65 | 54 | 44.71 | 44.95 | 10.74 | 37-51.3 |
| CD3+CD4+ count (cells*10<sup>3</sup>/μL) | 0.5-1.4 | 54 | 0.54 | 0.43 | 0.38 | 0.27-0.69 |
| CD3+CD8+ % | 12-40 | 54 | 23.36 | 24.15 | 9.82 | 15.6-30.5 |
| CD4+CD8+ count (cells*10<sup>3</sup>/μL) | 0.25-1 | 54 | 0.28 | 0.20 | 0.22 | 0.12-0.36 |
| CD19+ % | 5-20 | 51 | 12.39 | 11.50 | 6.88 | 8-14.83 |
| CD19+ count (cells*10<sup>3</sup>/μL) | 0.1-0.5 | 51 | 0.13 | 0.10 | 0.09 | 0.06-0.20 |
| Natural Killer % | 5-20 | 51 | 16.06 | 15.00 | 9.09 | 8-20.6 |
| Natural Killer count (cells*10<sup>3</sup>/μL) | 0.5-5 | 51 | 0.17 | 0.14 | 0.12 | 0.08-0.20 |
| Immunoglobulin G (mg/dL) | 650-1600 | 19 | 961.6 | 933.0 | 131.3 | 885-1006 |
| Immunoglobulin A (mg/dL) | 40-350 | 19 | 230.9 | 223.0 | 72.3 | 178-248 |
| Immunoglobulin M (mg/dL) | 50-300 | 19 | 103.1 | 90.0 | 39.8 | 72-129 |

Laboratory data at discharge
|  |  |  |  |  |  |  |
| --- | --- | --- | --- | --- | --- | --- |
| IL-6 (pg/mL) | <4.4 | 114 | 100.46 | 8.84 | 721.15 | 3.9-21.77 |
| CRP (mg/L) | <10 | 286 | 30.75 | 13.72 | 45.52 | 5-37.7 |
| ferritin (ng/mL) | 20-250 | 197 | 819.80 | 633 | 865.41 | 290-1163 |
| D- dimer (μg/L) | <500 | 256 | 3264.93 | 590.5 | 34427.29 | 366-1127 |
| LDH (U/L) | 120-246 | 257 | 272.76 | 234 | 312.86 | 195-280 |
| days from admission to discharge |  | 129 | 10.83 | 10 | 6.55 | 7-14 |
| Leucocyte count (cells*10<sup>3</sup>/μL) | 4-12.4 | 309 | 7.22 | 6.38 | 3.39 | 4.96-8.28 |
| Neutrophil count (cells*10<sup>3</sup>/μL) | 1.9-8 | 309 | 4.93 | 4.04 | 3.35 | 2.92-5.8 |
| Lymphocyte count (cells*10<sup>3</sup>/μL) | 0.9-5 | 309 | 1.52 | 1.43 | 0.76 | 1-1.9 |
| Lymphocyte % | 19-48 | 309 | 23.77 | 24 | 11.77 | 14.8-31.6 |
| CD3+CD4+ % | 25-65 | 13 | 49.86 | 54 | 14.69 | 49-58.24 |
| CD3+CD8+ % | 12-40 | 13 | 19.88 | 20 | 10.42 | 10-29.27 |
| CD19 % | 5-20 | 13 | 16.67 | 10.86 | 21.12 | 7.9-15 |
| Natural Killers % | 5-20 | 13 | 11.64 | 11 | 5.70 | 9-15 |

Abbreviations: RASB<sup>a</sup>, renin-angiotensin system blockers; Ref.v<sup>b</sup>, reference values; SD<sup>c</sup>, standard deviation; IQR<sup>d</sup>, interquartile range; IL-6<sup>e</sup>, interleukin-6; CRP<sup>f</sup>, C-reactive protein; LDH<sup>g</sup>, lactate dehydrogenase.

**Table 2.**
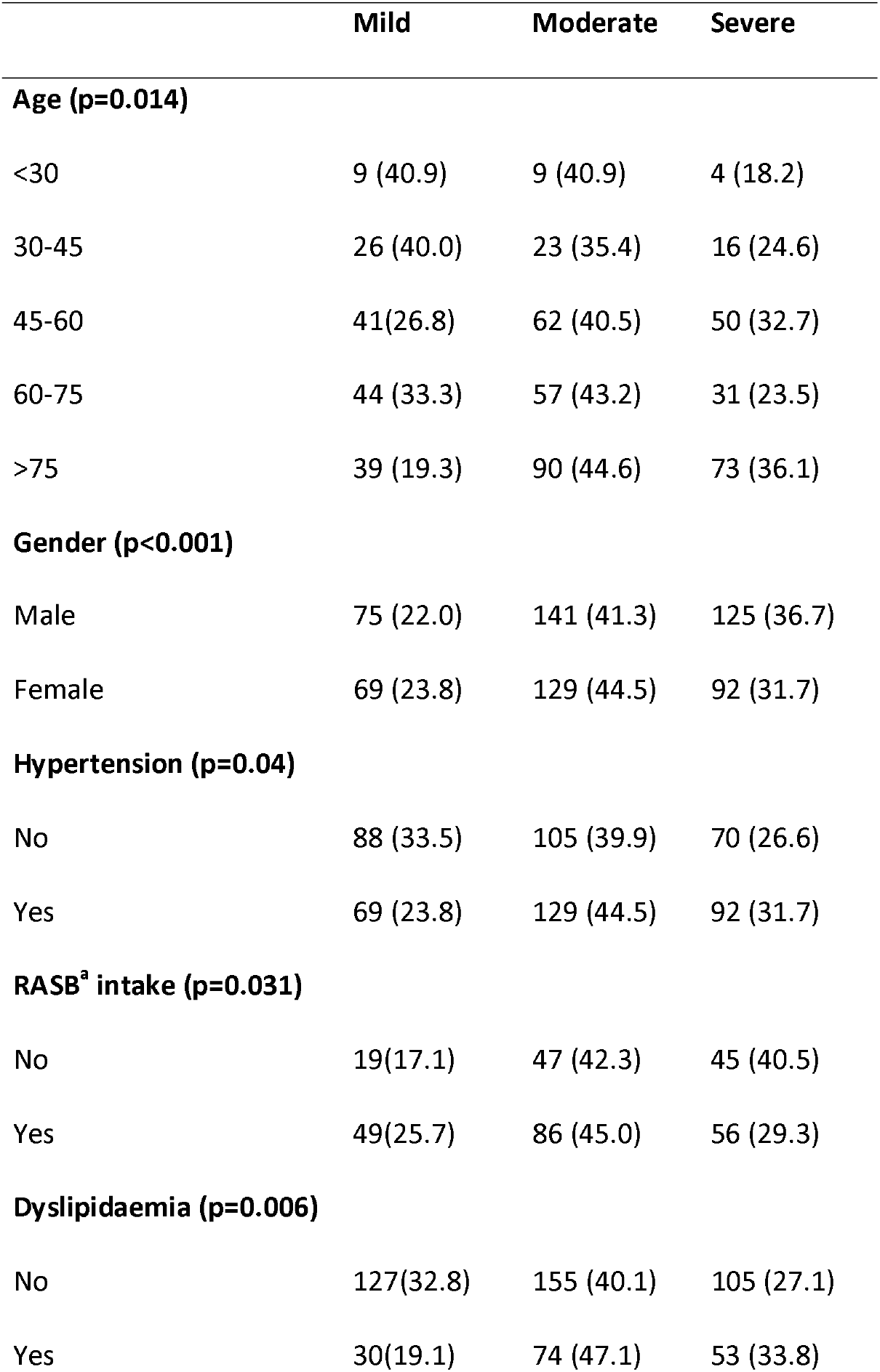
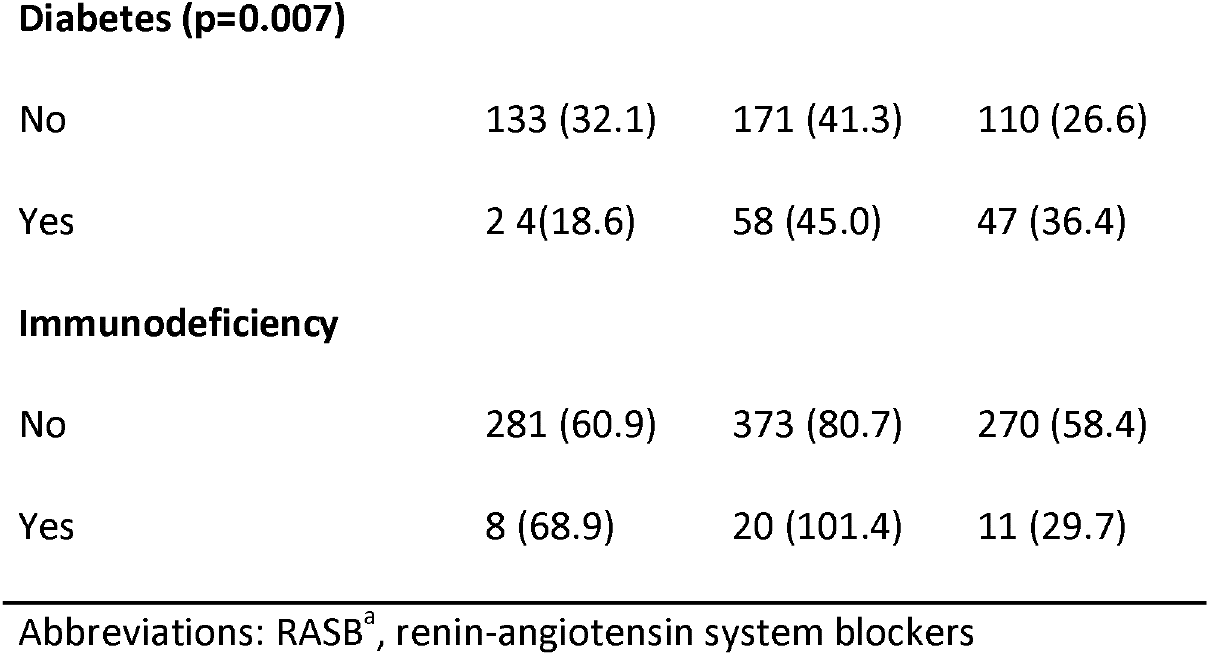
Risk Factors by severity.

|  | **Mild** | **Moderate** | **Severe** |
| --- | --- | --- | --- |
| **Age (p=0.014)** |  |  |  |
| <30 | 9 (40.9) | 9 (40.9) | 4 (18.2) |
| 30-45 | 26 (40.0) | 23 (35.4) | 16 (24.6) |
| 45-60 | 41(26.8) | 62 (40.5) | 50 (32.7) |
| 60-75 | 44 (33.3) | 57 (43.2) | 31 (23.5) |
| >75 | 39 (19.3) | 90 (44.6) | 73 (36.1) |
| **Gender (p<0.001)** |  |  |  |
| Male | 75 (22.0) | 141 (41.3) | 125 (36.7) |
| Female | 69 (23.8) | 129 (44.5) | 92 (31.7) |
| **Hypertension (p=0.04)** |  |  |  |
| No | 88 (33.5) | 105 (39.9) | 70 (26.6) |
| Yes | 69 (23.8) | 129 (44.5) | 92 (31.7) |
| **RASB<sup>3</sup> intake (p=0.031)** |  |  |  |
| No | 19(17.1) | 47 (42.3) | 45 (40.5) |
| Yes | 49(25.7) | 86 (45.0) | 56 (29.3) |
| **Dyslipidaemia (p=0.006)** |  |  |  |
| No | 127(32.8) | 155 (40.1) | 105 (27.1) |
| Yes | 30(19.1) | 74 (47.1) | 53 (33.8) |
| **Diabetes (p=0.007)** |  |  |  |
| No | 133 (32.1) | 171 (41.3) | 110 (26.6) |
| Yes | 24 (18.6) | 58 (45.0) | 47 (36.4) |
| **Immunodeficiency** |  |  |  |
| No | 281 (60.9) | 373 (80.7) | 270 (58.4) |
| Yes | 8 (68.9) | 20 (101.4) | 11 (29.7) |

Abbreviations: RASB<sup>a</sup>, renin-angiotensin system blockers

Fifty nine percent (59.4%) of cases were male. Ages in our cohort ranged from 18 to 98 years, with a mean of 63.2 years (SD 16.4). Concerning comorbidities, 52.4% were hypertensive, 63.2% of them were treated with blockers of renin-angiotensin system (RABs); twenty-eight percent 28.9% had dyslipidaemia and 23.8% suffered diabetes. Immunodeficiency diseases were most often secondary to other processes, such as a transplantation or chemotherapy treatment, these cases accounted for 8.2% (n=39) as seen in Table 1.

Both moderate and severe courses were found to be significantly associated with older age (p= 0.014), male gender (p<0.001), dyslipidaemia (p=0.006), hypertension (p=0.04) and diabetes (p=0.007). Severe cases over the age of 75 accounted for a 36.1%. The use of renin-angiotensin system blockers by hypertensive patients was associated, (p=0.031), with mild or moderate outcomes (Table 2).

Once at hospital, 84.8% of inpatients received antibiotics; the most commonly prescribed ones were azithromycin combinations (71.3%); those treated with antimalarial drugs accounted for 72.6% and 66% received antivirals, being litonavir/ritonavir the most widely used. Around one half, 51% of cases, received combined therapy with antibiotics, antimalarials and antivirals (commonly named triple therapy). Immunosuppressant drugs were used in 18.7% cases. Anti-cytokine therapy was used in 8.6%, mostly anti-IL-6R (Tocilizumab), and 17.7% were treated with interferons α or β (3 (Table 1).

On admission, means of laboratory parameters, IL-6, CRP, ferritin, D-dimer, LDH, leukocyte and neutrophil counts, were above reference ranges, opposite to lymphocyte subsets’ counts (Table 1). Higher severity was significantly associated with higher IL-6, CRP, ferritin, D-dimer, LDH, leukocyte and neutrophil counts, but lower lymphocyte percent (Supplementary Table 1). Mean values of lymphocyte subpopulations percentages (n=54) were within normality ranges. CD8 Lymphocyte count was found to be significantly higher in mild cases, a similar trend was found for CD4, CD8, CD19 and NK percentages. IgG and IgM values were as well inversely related to severity (Supplementary Table 1).

At discharge, IL-6, ferritin, D-dimer, LDH, leukocyte and neutrophil counts remained significantly higher within severe cases as compared to mild or moderate ones. Lymphocyte count turned significantly higher in severe cases as compared to mild or moderate (Supplementary Table 1). CRP values at discharge were nearly in range regardless of severity. Notwithstanding, when comparing laboratory data at discharge with those on admission, an overall return to reference ranges of most parameters was observed, with lower mean values of CRP, LDH and IL-6 and higher mean values of percentage and absolute lymphocyte count. D-dimer and ferritin still showed values above reference ranges with no significant differences as compared to these upon arrival.

Most differences among groups were kept as regarding aging (Figure 1). It could be evidenced that lymphopenia and increased IL-6 remained significant regarding severity in all age groups but in patients under 30. CD8 population differences (both in absolute count and percent) were significant only as regarding 45-60 group (the largest one). Lymphocyte count decrease –that was seen as a global change-was only evidenced for 30-45 and 45-60 age ranges. NK % was higher in milder cases within older individuals (60-75 and <75). Severity-related decreases of CD4 (p 0.007) and CD8 (p 0.008) lymphocytes were evidenced just in males (Figure 2). Hypertension, dyslipidaemia and diabetes grew more frequent with age (p<0.001, Table 3). These four risk factors showed strong interferences (Figure 3). Nevertheless, a predictive model couldn’t be proposed due to frequent missing values.

**Figure 1.**
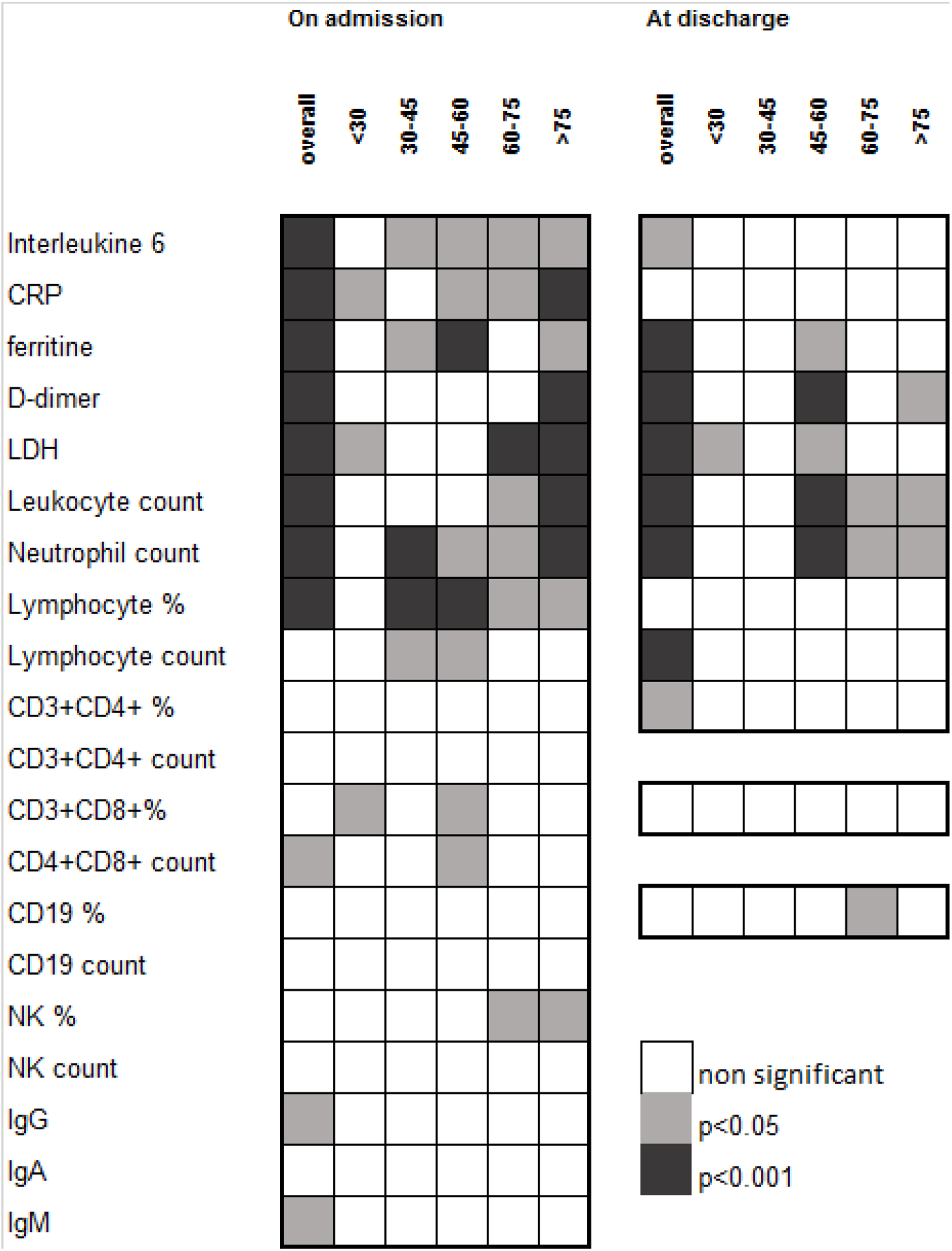
Severity related changes of laboratory parameters by age. Pearson’s Chi Squared p-values. Abbreviations: CRP, C-reactive protein; LDH, lactate dehydrogenase, NK, Natural Killers. IgG, immunoglobulin G; IgA, immunoglobulin A; IgM, immunoglobulin M.

**Figure 2.**
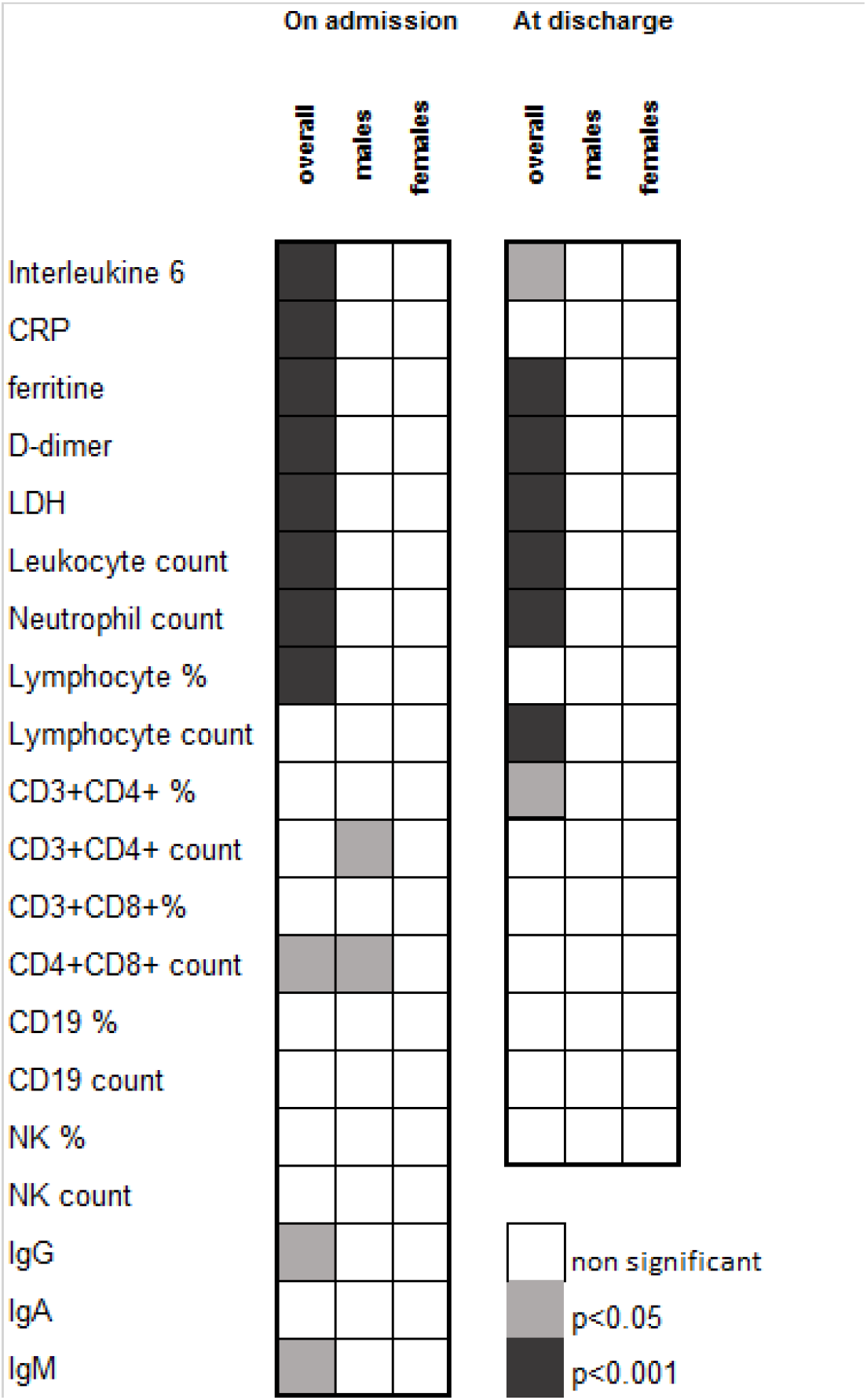
Severity related changes of laboratory parameters by gender. Pearson’s Chi Squared p-values Abbreviations: CRP, C-reactive protein; LDH, lactate dehydrogenase, NK, Natural Killers. IgG, immunoglobulin G; IgA, immunoglobulin A; IgM, immunoglobulin M.

**Table 3.**
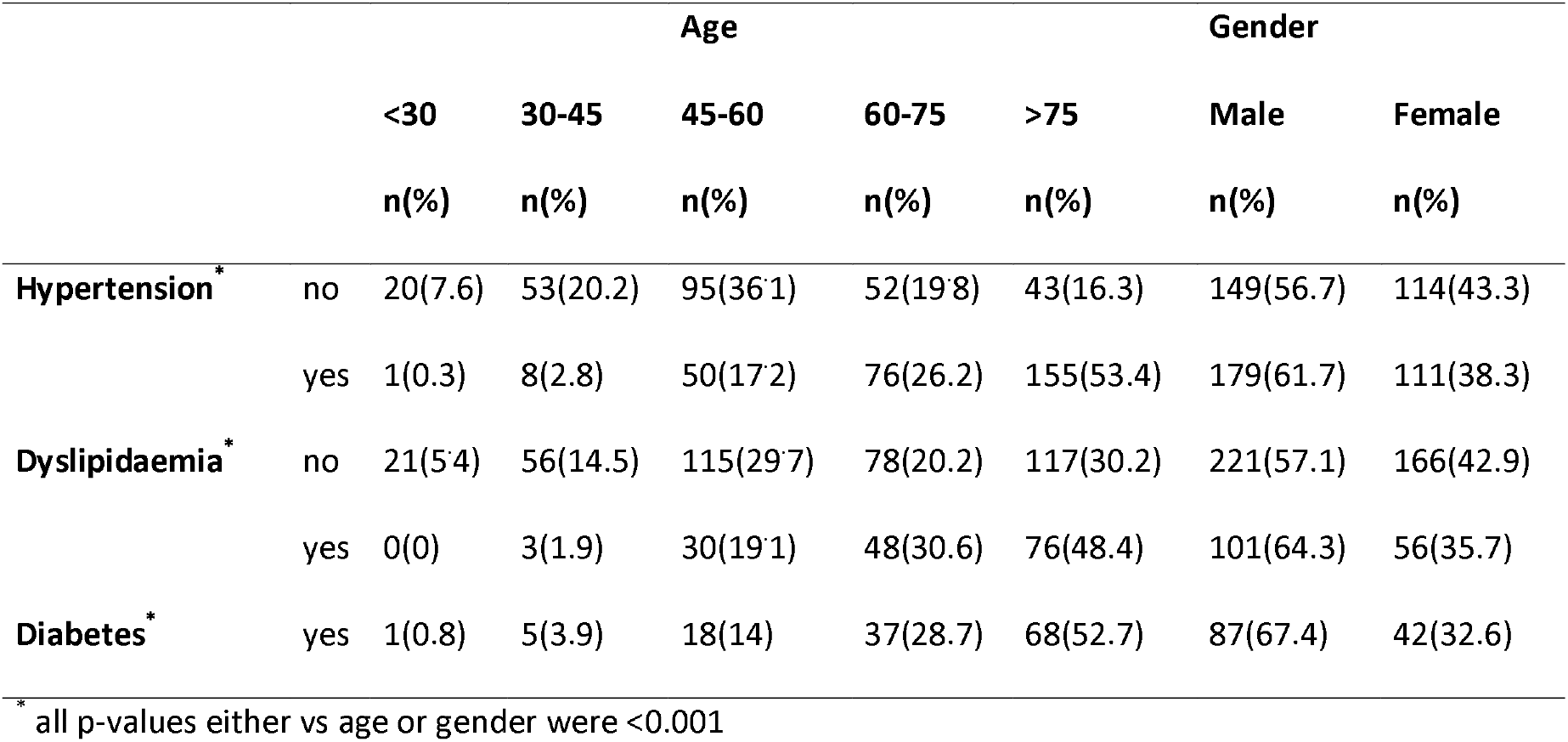
Influence of age and gender on comorbidities

**Figure 3.**
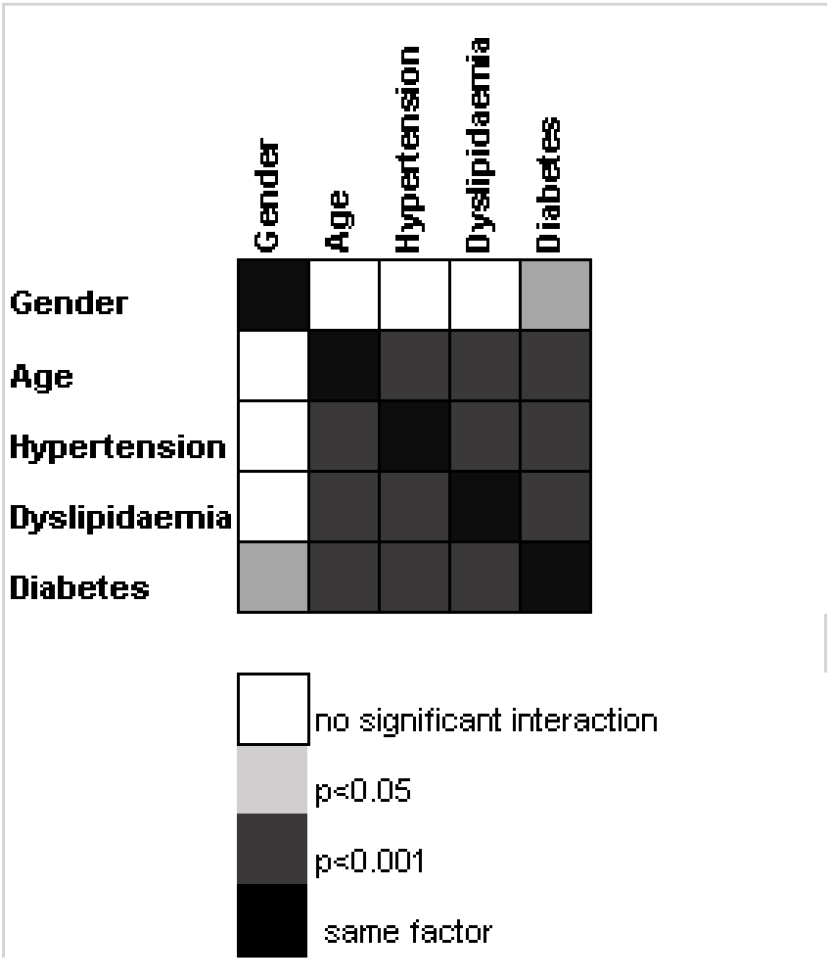
Severity factor interactions. Pearson’s Chi Squared p-values

## DISCUSSION

COVID-19 pandemic has become particularly virulent in Mediterranean countries such as Spain, both in terms of number of affected people and fatalities. This is the first report on Spanish COVID-19 inpatients; our aim was to outline disease’s demographic characteristics, risk factors and laboratory parameters, in relationship to disease severity.

In our series, 27.7% of patients showed a mild course, 42% a moderate course and 30.3% a severe one. Several works analyse severity in COVID-19 inpatients, almost all from Chinese hospitals. Those, including two multicentre studies, show a presentation in which the most severe cases ranged from 16% to 26% [16-19], except for the study of Zhou et al. [20], where critical cases reached 28%.

It has been reported elsewhere in papers on risk factors affecting the course of the disease, that older patients or those with at least one previous comorbidity had a worse prognosis [16,19-21], There are, however, remarkable differences within these studies. Spanish inpatients were much older than those in previously reported cohorts, with an average age of 63.2 years. Previous literature, including one meta-analysis and two multicentre studies, reports ages ranging from 36 to 58 [16-20,22-24], On the other hand, Grasselli et al. [25], in a multicentre Italian study focusing on patients admitted to the ICU, reports an average age of 63, similar to that in our cohort.

In SARS-CoV-2 infection, the number of paediatric patients is lower with milder symptoms and better prognosis [26] as compared to adults. This fact highlights the possible effect of immunosenescence on the course of the disease [27], Immunosenescence refers to the age-associated decline of the immune system, affecting both innate and adaptive immunity. Aging is associated with a greater susceptibility to infections [28]. People over 60 represent 11% of the worldwide population and they are expected to reach 22% by 2050 [28]. In Spain, to date, 19.4% of the population is over 65 [29].

Possibly due to ageing, it can be observed that frequencies for comorbidities such as hypertension or diabetes, were higher in our series than those reported in previous studies [16,17, 19-24, 30] and higher than the diabetes prevalence among Spanish adult population (23.5% vs 13.8%) [31]. Notwithstanding, the prevalence of hypertension mirrored those of the general adult Spanish population (overall, 42.6%; people over 60, 75.4%) [32], Not only SARSCoV2, but most human coronaviruses, strike harder the elderly and those with underlying comorbidities [33], probably as a consequence of a worse immune response control.

As in previous studies, patients were more frequently males [19, 20, 22, 24], This fact is even more noteworthy, considering that the percentage of men in the Spanish population over 50 years of age is 41.7% [29], under 59.4% in our COVID-19 cohort. In addition to remark, male gender was associated with severity. There is an Italian report of patients admitted to ICU, with a percentage of males reaching 80% [25] that would go in the same line of our observed gender effect on severity.

Concerning laboratory parameters, our findings were comparable to those reported in previous studies, with increases of acute phase reactants (CRP, D-dimer, LDH, ferritin) growing with severity and decreasing when the evolution of patients was favourable [19, 20, 22, 30]. Particularly striking was the evolution of the CRP, which was almost within reference range at discharge.

Several publications have focused on immunological markers in COVID-19 [19, 20, 30]. The most extensive one is the work of Diao et al. [14], which analyses the secretory profile of inflammatory cytokines, lymphocyte populations and their relationship to disease severity in 499 patients. The authors found an increase in pro-inflammatory cytokines inversely correlated to T-lymphocyte populations. This immune profile was as well related to the severity of the disease. CD4, CD8 and IL-6 are reported to covariate at least in mild cases [34] and behave like in our series, where an increase in IL-6 and a decrease in both total lymphocytes and lymphocyte populations could be seen. Once again, these changes were greater, the more severe the condition. However, in our study almost any lymphocyte population: CD4, CD8, CD19 and NK were below reference ranges upon arrival and were even more markedly decreased in severe cases, in spite of differences being only significant for CD8 population as regarding overall data.

Since the seminal publication of Lei Fang et al. on the possible involvement of renin-angiotensin system blockers in SARS-CoV-2 infection [10] just two months ago, there has been much ado about. No sooner had the scientific community realized its foreseeable impact, they began to take sides with articles both for and against the hypothesis [35-38]. ACE2 molecules are the pathway used by SARS-CoV-2 to enter the cell [39]. RASBs indirectly increase the expression and secretion to the extracellular medium of ACE2 in various cell types, including airway alveolar epithelial cells [35]. Therefore, RASBs use might enhance either the entry of the virus or its blockade preventing it from infecting the cell [36]. Additionally, the expression of ACE2 is associated with positive effects on lung homeostasis, those could be beneficial for tissue recovery from the damage caused by SARS-CoV-2 infection [11, 36]. ACE2 expression is reported to be related to age and sex. It would be highest in children and young women, decreasing with ageing, and would be lowered due to chronic disease comorbidities, including diabetes and hypertension [40]. ACE2 will inversely correlate severity and poor outcomes. Most literature for or against the role of the use of RASB consist mainly of theoretical positioning based on the knowledge of these drugs’ physiological actions. There are limited original studies analysing RASB intake effect on COVID-19. Tedeschi et al. [41], in order to elucidate whether RASB treatment had an impact on COVID-19 mortality, analysed 311 hypertensive patients hospitalized in 10 Italian centres. At multivariate Cox regression analysis of intra-hospital mortality, the use of RASB was not found to be associated with outcome. Moreover, Chen et al. [30] reported 113 hypertensive patients, 33 (29.2%) that were on RASB treatment, 87.9% of whom had a moderate course of the disease and 12.2% a severe or critical course. In our series, out of 290 hypertensive patients, 190 (67.4%) were taking RASB; a feature comparable to the intake of these compounds by the Spanish hypertensive population [32], Those patients treated with RASB drugs had a milder course of the disease (OR 0.61, 95% CI [0.37-0.99]; p 0.03). The present study has two major limitations. The firs one is derived from its retrospective design. As we are reporting on the very first cases of the disease in Spain, several immunological parameters of interest were not systematically tested. The other constraint is the short follow-up period of patients, which limits the possibility to have a full track of those who were still in hospital by data collection deadline. Consequently, the relationship among other variables such as treatment or other outcomes such as death, have not been here analysed.

## CONCLUSIONS

To summarize, our patients are older and developed more often severe COVID-19 condition than the previously reported cohorts. Age has emerged as a crucial factor in our series. Age is as well one of the major determinants for all other COVID-19 risk comorbidities, such as hypertension, diabetes, or dyslipidaemia. Immunosenescence might be a suitable explanation for the immune overwhelming observed in the severest cases. Regarding not only our series but other worldwide ones, the effectors of the immune system are hampered as severity increases. Adaptive immunity has been suggested to be disabled by SARS CoV2, that feature is so named exhausted immunity. This exhaustion would be coupled with a huge ineffective and almost deleterious innate response.

Further studies specifically aimed at assessing the immune system status in SARS-CoV-2 infected patients should be carried out to support immunosenescence hypothesis. Nevertheless, it is clear from data that elderly are at special COVID-19 risk and should be therefore paid special attention by public health services.

## METHODS

### Aim, design and setting of the study

The aim of this study was to obtain, within the shortest possible time, a reliable snapshot of the demographic and clinical characteristics of COVID-19 patients admitted to Spanish hospitals along the first month of the pandemic and to reveal risk factors regarding severity. So, a retrospective observational multicentre analysis was performed in 19 Spanish hospitals.

### Participants

The study population comprised the first consecutive set of SARS-CoV-2 infected inpatients, microbiologically confirmed by positive PCR test, during the second half of March 2020. Cases were tracked for a three-week follow-up period from admission to discharge. A minimum sample size of 20 patients was considered for every hospital. In some cases, this minimum was exceeded. A total of 642 medical records of individuals over 18 years old, from 19 Spanish hospitals were initially reviewed. After data quality assessment, 574 patients were finally included in the analyses. Participants were stratified into three severity groups before analysis according to the following clinical criteria:

- Mild: individuals whose clinical symptoms were mild with no abnormal radiological findings
- Moderate: cases with confirmed pneumonia that was not considered severe
- Severe: when at least one of the following criteria was met: acute respiratory distress, shock, admission to the intensive care unit (ICU), or the process was so considered by the physician in charge. Any “exitus” was as well classified as severe.

### Data collection

All data were extracted from electronic medical records. The collection form included demographic, epidemiological and clinical data: age, sex, history of diabetes mellitus (DM), dyslipidaemia, hypertension (HTA), renin-angiotensin system blocker intake (RASB), COVID-19 severity, time from onset to diagnosis, laboratory data on admission and discharge, treatment, and outcome. At the end of data collection, some patients were still in hospital. In these cases, laboratory data at discharge could not be provided.

### Laboratory data

Major laboratory markers were extracted from medical records at admission and discharge. Routine blood examinations included leukocyte, neutrophil and lymphocyte count (cells* 10^^^3/μL) and lymphocyte percentage. Serum biochemical tests recorded were ferritin (μg/L), lactate dehydrogenase (LDH, U/L), C-reactive protein (CRP) and D-dimer (μg/L). Immunological tests recorded were interleukin-6 (IL-6, pg/mL), Lymphocyte population count (cells*10^^^3/μL) and percentage by flow cytometry, immunoglobulins IgG, IgA and IgM (mg/dL).

### Statistical analysis

Demographic and clinical characteristics of patients were expressed as their mean and standard deviation (SD); when not adjusting to a normal distribution, median was used to represent non-parametrical data for continuous variables and frequency distributions are reported for categorical variables. Age was analysed both, as continuous and categorical variable; in the latter case was recoded into 5 groups: <30, 30-45, 45-60, 60-75, >75

Kolmogorov-Smirnov test was performed on each continuous variable to contrast normality. To analyse the overall differences between the three groups: mild, moderate, and severe type, ANOVA was tested on variables with normal distribution and n>30 (age, % and CD4 lymphocyte count, % of CD8 lymphocytes, % of CD19 lymphocytes and % of NK). To analyse severity relationships of non-parametric or n<30 variables, a Kruskal-Wallis test was used. A t-test was performed to contrast damage-free discharge for normal variables and a Mann Whitney test for non-parametric measures. To contrast the Ho of independence within categorical variables, Pearson’s Chi-square and Fisher’s exact test were used. To compare values of recurrent parameters measured in a same individual at admission and discharge, Wilcoxon test for paired data was performed.

## Data Availability

Data referred are available as an anonymized data base under requirements approbed by our ethics committee

**Abbreviations**
ACE2: Angiotensin-converting enzyme 2
RASB: renin-angiotensin system blockers
LDH: lactate dehydrogenase
CRP: C-reactive protein
IL-6: interleukin-6
IgG: immunoglobulin G
IgA: immunoglobulin A
IgM: immunoglobulin M
PCR: polymerase chain reaction
SD: standard deviation
IQR: interquartile range
NK: Natural Killers

## DECLARATIONS

### Ethics approval

This study was conducted according with national regulations, institutional policies and in the tenets of the Helsinki Declaration. This study was approved, with the Valladolid Health Area Drug Research Ethics Committee acting as the main committee, in a meeting held on March 31, 2020 and with the reference number “PI 20-172-NO-HCU”. Moreover, it was approved by each of the local institutional Ethics Committee of the 19 hospitals involved.

### Availability of data and materials

Data collected for the study, including fully anonymized participant data, are available to others. Data available include fully anonymized participant data and data dictionary. Related documents are available from the date of publications henceforth: study protocol, statistical analysis, and approval of Ethical Board. These documents are available from the date of publications henceforth at email address

Data will be shared after approval of proposals by the Valladolid Este Ethical Committee.

### Authors’ contributions

AJ and MCM conceived the idea for this study, designed the protocol, analysed the data and drafted the manuscript. The remaining authors collected the data and assessed for data quality. All authors provided critical revisions and approved the final version of the manuscript.

## Acknowledgements

The authors thank to all caregivers who indirectly contributed to this multicentre study. They also thank Spanish Immunology Society for its support and endorsement

